# Early life multidimensional disadvantage of South Australian children: a whole-population linked data study

**DOI:** 10.64898/2026.06.03.26354860

**Authors:** Anna Kalamkarian, Rhiannon M. Pilkington, John Lynch, Murthy N Mittinty, Catia Malvaso, Katherine Hawkins, Henry Pharo, Kerry Beck, Catherine R. Chittleborough

**Affiliations:** School of Public Health, Adelaide University, Adelaide, Australia; Population Health Sciences, University of Bristol, Bristol, United Kingdom; College of Medicine and Public Health, Flinders University, Adelaide, Australia; School of Psychology, Adelaide University, Adelaide, Australia; Robinson Research Institute, Adelaide University, Adelaide, Australia; Department of Human Services, Government of South Australia, Adelaide, Australia

**Keywords:** Multidimensional disadvantage, early childhood, linked administrative data

## Abstract

**Background:** Whole-population linked administrative data platforms provide an opportunity to generate evidence on early life multidimensional disadvantage to inform resourcing and service provision to families with complex needs.

**Methods:** We used individual-level de-identified data from nine administrative data sources included in the Better Evidence Better Outcomes Linked Data (BEBOLD) platform. The population included all children born in South Australia between 2004-2011 (n=143,083), and their parents. We described the prevalence and distribution of multiple disadvantages affecting children from the 12 months before birth to age 5. Eleven domains of parental disadvantage were created: economic, education, access to services, mental health, substance misuse, smoking during pregnancy, domestic and family violence, health, child protection contact, justice system contact, and death. We investigated the concordance of our measure with an area-level socioeconomic measure used in government reporting.

**Results:** One in two children (48%) were exposed to at least one disadvantage domain, and one in seven (14%) were exposed to three or more domains before age five. Economic disadvantage was most prevalent, affecting one in four (27%) children, of which 75% were exposed to additional forms of disadvantage. Substance misuse, domestic and family violence, and justice system contact were the least likely domains to occur in isolation. Only 54.4% who experienced five or more disadvantage domains were classified in the area-level socioeconomic measure’s ‘most disadvantaged’ quintile.

**Conclusion:** Early life exposure to parental disadvantage can be highly multidimensional. Measurement across different systems is important for informing coordinated service provision for families with complex needs.

**What is already known on this topic:** Children can experience a range of disadvantages in early life which can influence their health, development and long-term outcomes.

**What this study adds:** Linked administrative data makes it possible to observe co-occurrence of disadvantages across multiple service systems, beyond what is captured by area-level socioeconomic measures. This approach is particularly valuable for disadvantage measurement in early childhood, when children rely on parents and systems to represent their disadvantage.

**How this study might affect research, practice or policy:** This study highlights the potential of linked administrative data to identify parents’ overlaps in service system use over time. These insights can inform the resourcing and coordination of supportive services for families with complex needs.

## Introduction

In 1864, William Farr identified how many children’s lives were unnecessarily lost in England as a consequence of residing in areas deemed to be unhealthy.^(1)^ Today, the health and wellbeing of children is still shaped by the environments they are born into, which are influenced by parents’ economic, social and health circumstances. Parental exposure to disadvantage prior to and in the years after their child’s birth is associated with an increased risk of a range of poor health and development outcomes in a child’s life.^(2)^ While developing in-utero, children may be at risk of poor health outcomes through exposure to disadvantage in the form of, for example, smoking during pregnancy,^(3)^ maternal substance misuse,^(4)^ or violence directed at the pregnant woman.^(5)^ Household income and material deprivation have direct impacts on children’s development, housing and access to resources.^(6)^ Children may also be adversely impacted by circumstances that reduce parental capacity, including parent’s mental and physical health, substance misuse, disability, their own experiences of child maltreatment, imprisonment, discrimination, and experience of domestic and family violence.^(7, 8)^

Globally, there is strong socio-political commitment in reducing early childhood disadvantage and ensuring all children have opportunity to thrive into adulthood.^(9)^ The early years, from pre-birth until starting school, are recognised as a crucial time when early support can improve long-term health and development outcomes.^(10)^ However, support provision is constrained by measurement challenges in determining how many children require support and in resourcing that support accordingly.

In child health research, the concept of multiple disadvantage gained scientific and policy traction since the development of the Adverse Childhood Experiences (ACEs) scale in the late 1990s.^(11)^ However, screening for ACEs has been subject to criticism that it relies on limited recall of early childhood experiences,^(12)^ is rarely linked to prevention and provision of early support,^(13)^ and often excludes measurement of socioeconomic circumstances despite their central role in shaping children’s outcomes.^(14)^ One challenge in measuring early childhood disadvantage is that young children rely on parents or systems to represent their disadvantage, unlike later years when self-report becomes possible. Other multidimensional disadvantage constructs have been created to measure childhood disadvantages, of which, some studies centre predominantly on child-specific deprivations,^(15, 16)^ while others include more parent-specific disadvantages.^(17, 18)^

Parents’ experiences of disadvantage can provide a useful proxy for early childhood disadvantage, based on the assumptions that disadvantages have the potential to reduce parenting capacity^(7)^ or create intergenerational cycles of disadvantage.^(19)^ Understanding the types of disadvantage parents experience is therefore useful for informing the kinds of supports families require. Inquiries into child deaths have frequently identified a failure of agencies to work together to timely provide services to families.^(20, 21)^ Information systems and service provision often operate in silos, resulting in missed opportunities to address disadvantages before poor outcomes occur.^(22)^ A key barrier to designing coordinated responses is limited population-level evidence of the disadvantage co-occurrence throughout early childhood.

Ideally, having population-wide, routinely collected data on disadvantages of children and their parents could support routine planning and resourcing of services. In the absence of such individual-level information, geographical area-level measures have been used to estimate the extent of disadvantage in the population and their effects on children’s outcomes; example measures include the Australian Socio-Economic Indexes for Areas (SEIFA),^(23)^ Indices of Disadvantage in the UK,^(24)^ and the Area Deprivation Index in the US.^(25)^ These indices use administrative or census data on education, income, employment and other domains to produce scores that rank areas based on their relative socioeconomic advantage and disadvantage. While providing insight on socioeconomic disadvantage within a community, area-level measures are not able to capture the diversity of individual disadvantage, as not everyone in a geographical area experiences the same degree or types of disadvantage.^(26)^ Additionally, these measures rarely include indicators informative for understanding families’ complex needs, such as mental health or substance misuse exposure.^(8)^

Using de-identified linked administrative data from the Better Evidence Better Outcomes Linked Data (BEBOLD) platform, we quantified children’s exposure to mother and co-parent disadvantage, from the year before birth up to age 5. Our objective was to describe the prevalence and co-occurrence of disadvantage domains from before birth to age five, and to examine the concordance between our multidimensional measure of disadvantage and the SEIFA Index of Relative Socioeconomic Advantage and Disadvantage (IRSAD), an area-level socioeconomic measure used in Australian government reporting.

This study is deliberately descriptive to quantify the prevalence and co-occurrence of early childhood disadvantage captured across multiple service systems. Whole-of-population linked administrative data on children and their parents provides an opportunity to overcome measurement challenges by capturing children’s individual-level exposure to disadvantage through parents’ interactions with service systems at each age of the child.

## Methods

### Data Sources

Data were sourced from the BEBOLD platform including de-identified linked administrative data of South Australian (SA) children born 1991 onwards, and their parents.^(27)^ Data linkage was conducted by SA NT DataLink, a third-party accredited linkage authority, operating under strict privacy protecting practices.^(28)^ SA NT DataLink uses a probabilistic linkage algorithm to match individuals across the datasets using information including date of birth, name, sex and address. False linkage rates have been estimated to be 0.4% to 0.8%.^(29)^

The study population included children with a registered birth in SA between 1^st^ of July 2004 and 31^st^ of December 2011 (n=143,378), with information available for all mothers and 139,534 co-parents. The unit of analysis was the child and, as parents can have multiple children, there were 99,696 unique mothers and 97,592 unique co-parents. These birth cohorts were chosen so that information on all indicators was available from one year before each child’s birth, and for five years after birth up to 2016.

Children with a death record were included in age-specific analyses until the year in which the death occurred. There were 143,083 children who survived until age 5 (Supplementary Figure 1).

### Disadvantage Indicators and Domains

Eleven domains of parents’ circumstances and experiences were included to capture the disadvantaged circumstances that children are born and grow in, until age 5: economic, education, access to services, mental health, substance misuse, domestic and family violence, health, smoking in pregnancy, child protection contact, contact with the justice system, and death. Each domain comprised a range of individual-level indicators denoting a form of constraint on parents’ availability or ability to meet children’s needs. An indicator was considered to meet the conditions of disadvantage if it: (1) had evidence of being detrimental to children’s outcomes, and (2) was potentially preventable through government policies or interventions.

Details on the definition and data sources for each indicator is provided in Supplementary Table 1 to Supplementary Table 11. Parental education, remoteness of area of residence, and parents’ history of child protection system contact were measured at the birth of the child and assumed to remain unchanged over the course of the child’s first five years of life. For the ‘access to services’ domain, sensitivity analyses in a subsample with data collected in the Australian Early Development Census (n=35,830) demonstrated that the indicator changed minimally over early life with only 2.1% of children misclassified by age 5 (Supplementary Table 12).

### Data analysis

Each child was classified as disadvantaged on a particular domain if they experienced at least one indicator within that domain. Age-specific prevalence was examined by counting whether the indicator or domain had occurred from the child’s birthday through to the day before they turned the next year older. A child was classified as ‘ever’ disadvantaged if they experienced the indicator or domain at any age from 12 months before their birth, up to their 5^th^ birthday.

The number of different disadvantage domains experienced at each age was calculated. There was a total of ten possible disadvantage domains that children could be exposed to at ages one to five, and up to eleven possible disadvantage domains in the 12 months before birth due to the smoking in pregnancy domain. For domains where information was measured over time, persistence of disadvantage domains was investigated by counting the number of years that the disadvantage domain occurred, with findings reported in Supplementary Table 13.

The proportion of children experiencing a disadvantage domain in isolation or in combination with one, two, three, or four or more other domains was calculated.

The proportion of children who experienced disadvantage on each domain and each possible combination of domains was illustrated using a “dashboard approach,”^(30)^ constructed from *UpSet* plots.^(31)^ Each *UpSet* plot was truncated to show intersections of the 20 most prevalent combinations of disadvantages among children who experienced at least one domain of disadvantage. The total prevalence of each domain was represented in the horizontal left bar plot, and intersections were shown in the vertical plot. Further *UpSet* plots visualised commonly occurring disadvantage domain combinations among children experiencing disadvantage in terms of parents’ mental health, substance use, domestic and family violence, and justice system contact, as combinations of these domains are dominant in literature concerning children’s welfare.^(7, 32, 33)^

The distribution of each disadvantage domain, and the number of disadvantage domains experienced, over the six-year time period between 12 months before birth to age 5 was examined across quintiles of area-level disadvantage according to the area of residence recorded at the child’s birth. The Socio-Economic Indexes For Areas (SEIFA) Index of Relative Socio-economic Advantage and Disadvantage (IRSAD) ranks small areas, according to the economic and social conditions of households within those areas using Census data.^(23)^ IRSAD is predominantly based on indicators of income, education, employment and housing, and does not include domains such as health, domestic and family violence, substance misuse, contact with child protection or justice systems. A low score indicates relatively greater disadvantage and lack of advantage. Children with missing information on IRSAD (n=112) were excluded from the comparison.

### Ethics

Ethics and Site-Specific Assessment (SSA) approval was granted from the South Australian Department of Health and Wellbeing Human Research Ethics Committee (2022/HRE00137).

## Results

### Disadvantage indicators and domains

Table 1 demonstrates that the prevalence of single indicators of disadvantage varied from 16.8% for an occupant of a household receiving rental bond assistance, to 0.5% who experienced a parental death. The most common disadvantage domain was economic (26.5%). Health disadvantage was the second most prevalent, with 1 in 5 children (20.5%) exposed to parental health disadvantage by age 5 (Table 1). The least prevalent disadvantages over the six-year period were domestic and family violence (2.3%), parental justice system contact (1.8%), or death of parents (0.5%). Further age-specific disadvantage indicators and breakdown of mother and co-parent contributions to each indicator are available in Supplementary Table 14 to Supplementary Table 29.

**Table 1:** Proportion of children who ever experienced disadvantage domains and indicators from 12 months before birth to age 5 (N=143,083)

|  | n | % | (95% CI) |
| --- | --- | --- | --- |
| **Economic - at least one of the following:** | **37,924** | **26.5** | **(26.3 – 26.7)** |
| Child (or their parent before birth) listed on application in household receiving private rental bond assistance | 23,970 | 16.8 | (16.6 – 16.9) |
| Child had school card at age of school entry | 18,111 | 12.7 | (12.5 – 12.8) |
| Child (or their parent before birth) listed in public housing or registered on public housing waitlist | 15,475 | 10.8 | (10.7 – 11.0) |
| Mother or co-parent had hospital admission related to unemployment or problems with income or housing | 1,352 | 0.9 | (0.8 – 0.9) |
| Child in jobless family | 11,739 | 8.1 | (8.0 – 8.3) |
| **Education** |  |  |  |
| Parents' highest level of education attained was Year 11 or equivalent or below | 9,118 | 6.4 | (6.2 – 6.5) |
| **Access to services** |  |  |  |
| Mother living in geographically remote or very remote area | 5,462 | 3.8 | (3.7 – 3.9) |
| **Mental health - at least one of the following:** | **18,248** | **12.8** | **(12.6 – 12.9)** |
| Mother or co-parent had hospital admission related to mental health | 14,922 | 10.4 | (10.3 – 10.6) |
| Mother or co-parent had ED presentation related to mental health | 8,769 | 6.1 | (6.0 – 6.3) |
| **Substance misuse - at least one of the following:** | **9,780** | **6.8** | **(6.7 – 6.9)** |
| Mother or co-parent had contact with DASSA services | 703 | 0.5 | (0.5 – 0.5) |
| Mother or co-parent had hospital admission related to substance misuse | 8,466 | 5.9 | (5.8 – 6.0) |
| Mother or co-parent had ED presentation related to substance misuse | 2,893 | 2.0 | (2.0 – 2.1) |
| **Domestic and family violence - at least one of the following:** | **3,346** | **2.3** | **(2.3 – 2.4)** |
| Mother or co-parent had hospital admission related to domestic and family violence | 858 | 0.6 | (0.6 – 0.6) |
| Domestic family violence concern in public housing | 2,660 | 1.9 | (1.8 – 1.9) |
| **Smoking in pregnancy** |  |  |  |
| Mother smoked during pregnancy | 23,652 | 16.5 | (16.3 – 16.7) |
| **Health - at least one of the following:** | **29,327** | **20.5** | **(20.3 – 20.7)** |
| Mother or co-parent ever had up to 4 or more hospitalisations within 12 months | 7,241 | 5.1 | (4.9 – 5.2) |
| Mother or co-parent ever stayed in hospital for over 8 days within 12 months | 17,382 | 12.2 | (12.0 – 12.3) |
| Mother or co-parent ever had up to 4 or more ED presentation within 12 months | 16,123 | 11.3 | (11.1 – 11.4) |
| **Child protection contact** |  |  |  |
| Mother or co-parent had CP substantiation or OOHC placement | 8,630 | 6.0 | (5.9 – 6.2) |
| **Justice system contact** |  |  |  |
| Mother or co-parent experienced imprisonment | 2,534 | 1.8 | (1.7 – 1.8) |
| **Death** |  |  |  |
| Child experienced death of parent | 752 | 0.5 | (0.5 – 0.6) |

NB 'Parent' refers to either mother or father

Table 2 shows the number of disadvantage domains experienced at each age. Overall, between the 12 months before birth up to age 5, one in two children (47.9%) had experienced one or more disadvantage domains, and 4.6% experienced five or more disadvantage domains. The highest exposure to disadvantage occurred in the 12 months prior to birth, with 36.6% experiencing one or more domains of disadvantage and 1.1% experiencing five or more domains, compared to 27.2% experiencing one or more domains, and 0.5% experiencing five or more domains at age 5. Age-specific prevalence of each disadvantage domain is provided in Supplementary Table 30.

**Table 2:** Number of disadvantage domains experienced at each age.

| Number of disadvantage domains | 12 months before birth (n=143,378) |  | Age 1 (143,180) |  | Age 2 (n=143,128) |  | Age 3 (n=143,108) |  | Age 4 (n=143,098) |  | Age 5 (n=143,083) |  | Overall from 12 months before birth to age 5 (n=143,083) |  |
| --- | --- | --- | --- | --- | --- | --- | --- | --- | --- | --- | --- | --- | --- | --- |
|  | n | % | n | % | n | % | n | % | n | % | n | % | n | % |
| None | 90,860 | 63.4 | 108,530 | 75.8 | 110,196 | 77.0 | 110,145 | 77.0 | 110,783 | 77.4 | 104,160 | 72.8 | 74,573 | 52.1 |
| 1 domain | 31,074 | 21.7 | 22,843 | 16.0 | 22,056 | 15.4 | 22,093 | 15.4 | 21,623 | 15.1 | 24,461 | 17.1 | 31,832 | 22.3 |
| 2 domains | 12,155 | 8.5 | 7,315 | 5.1 | 6,880 | 4.8 | 6,846 | 4.8 | 6,612 | 4.6 | 9,313 | 6.5 | 16,088 | 11.2 |
| 3 domains | 5,357 | 3.7 | 2,787 | 2.0 | 2,509 | 1.8 | 2,511 | 1.8 | 2,533 | 1.8 | 3,281 | 2.3 | 8,847 | 6.2 |
| 4 domains | 2,340 | 1.6 | 1,153 | 0.8 | 953 | 0.7 | 996 | 0.7 | 1,034 | 0.7 | 1,184 | 0.8 | 5,167 | 3.6 |
| 5 or more domains | 1,592 | 1.1 | 552 | 0.4 | 534 | 0.4 | 517 | 0.4 | 513 | 0.4 | 684 | 0.5 | 6,576 | 4.6 |

NB Smoking during pregnancy was only included in the age-specific analysis before birth and not at ages 1 to 5

Of the 68,510 children exposed to one or more disadvantage domains between 12 months before birth to age 5, 19.9% experienced disadvantage only before birth and 23.6% experienced disadvantage only after birth; therefore, the majority of children (56.5%) who were exposed to at least one disadvantage domain, were exposed both before and after birth [data not shown].

### Co-occurrence of disadvantage domains

There were 655 unique combinations of disadvantage domains, with the 20 most common combinations shown in Figure 1. Single disadvantage domains in isolation were most prevalent (14.5% for health, 14.0% for economic, 7.7% for smoking in pregnancy), followed by two domains in combination (4.9% economic and health, 4.6% economic and smoking in pregnancy). Of all 655 disadvantage combinations, more than half (55.4%) involved economic disadvantage (Figure 1). Table 3 shows only 25.3% of all children experiencing at least one disadvantage domain were exposed to economic disadvantage in isolation, with the remainder experiencing economic disadvantage alongside a combination of other disadvantages. The most common combination of five-dimensional disadvantage was the experience of economic, health, smoking in pregnancy, mental health and substance misuse disadvantage, which affected 1.3% of children, and a further 3.0% of children who experienced this combination alongside other disadvantages [data not shown].

**Figure 1.**
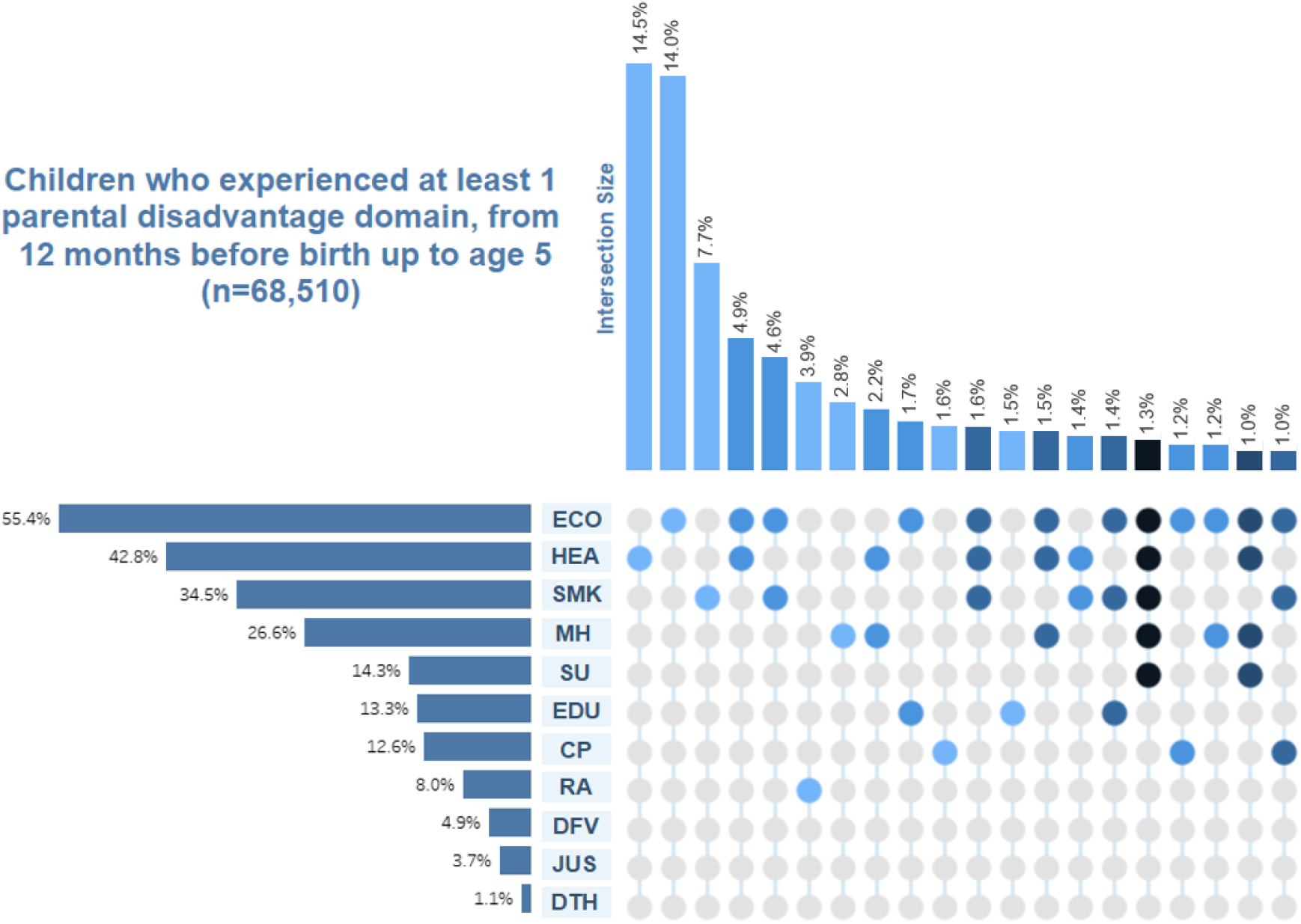
Prevalence of each disadvantage domain, and the 20 most prevalent combinations of disadvantage domains (out of total 655 combinations), for children who ever experienced at least one disadvantage domain from 12 months before birth to age 5 (n=68,510) Disadvantage domains: ECO = economic, HEA = health, SMK = smoking during pregnancy, MH = mental health, SU = substance misuse, EDU = education, CP = child protection contact, RA = access to services, DFV = domestic and family violence, JUS = justice system contact, DTH = death of parent

**Table 3:** Proportion of children experiencing each disadvantage domain in isolation or co-occurring with other disadvantage domains, from 12 months before birth up to age 5.

| Parental disadvantage domain | Domain occurred in isolation |  | Co-occurrence with 1 other domain |  | Co-occurrence with 2 other domains |  | Co-occurrence with 3 other domains |  | Co-occurrence with 4 or more other domains |  |
| --- | --- | --- | --- | --- | --- | --- | --- | --- | --- | --- |
|  | n | % | n | % | n | % | n | % | n | % |
| Economic (n=37,924) | 9,605 | 25.3 | 10,178 | 26.8 | 7,108 | 18.7 | 4,603 | 12.1 | 6,430 | 17.0 |
| Education<sup>(a)</sup> (n=9,118) | 1,015 | 11.1 | 1,925 | 21.1 | 2,118 | 23.2 | 1,450 | 15.9 | 2,610 | 28.6 |
| Access to services<sup>(a)</sup> (n=5,462) | 2,674 | 49.0 | 1,336 | 24.5 | 646 | 11.8 | 347 | 6.4 | 459 | 8.4 |
| Mental health (n=18,248) | 1,952 | 10.7 | 3,349 | 18.4 | 3,567 | 19.6 | 3,386 | 18.6 | 5,994 | 32.9 |
| Substance misuse (n=9,780) | 43 | 0.4 | 718 | 7.3 | 1,519 | 15.5 | 2,183 | 22.3 | 5,317 | 54.4 |
| Domestic and family violence (n=3,335) | 19 | 0.6 | 386 | 11.6 | 575 | 17.2 | 568 | 17.0 | 1,787 | 53.6 |
| Health (n=29,327) | 9,918 | 33.8 | 6,908 | 23.6 | 4,378 | 14.9 | 3,066 | 10.5 | 5,057 | 17.2 |
| Smoking in pregnancy (n=23,652) | 5,283 | 22.3 | 5,463 | 23.1 | 4,433 | 18.7 | 3,159 | 13.4 | 5,314 | 22.5 |
| Child protection contact<sup>(a)</sup> (n=8,630) | 1,115 | 12.9 | 1,511 | 17.5 | 1,720 | 19.9 | 1,407 | 16.3 | 2,877 | 33.3 |
| Justice System contact (n=2,534) | 104 | 4.1 | 220 | 8.7 | 344 | 13.6 | 402 | 15.9 | 1,464 | 57.8 |
| Death (n=752) | 104 | 13.8 | 182 | 24.2 | 133 | 17.7 | 97 | 12.9 | 236 | 31.4 |

<sup>(a)</sup> Indicators for 'education,' 'access to services' and 'child protection' domains were captured at one point in time and assumed to stay consistent over the duration of six-year study period

Figure 1 demonstrates that 1 in 3 (34.5%) children experiencing at least one disadvantage domain were exposed to pre-natal smoking, yet only 7.7% of children experienced this disadvantage in isolation. The access to services domain had the lowest co-occurrence levels; 8.0% of children who experienced at least one disadvantage domain lived in remote or very remote areas, and half of these children (3.9%) experienced no other disadvantage (Figure 1).

A small proportion of children were exposed to substance use, domestic and family violence, or justice system contact, but these disadvantage domains were extremely likely to co-occur with other disadvantage domains. Among children who experienced parents’ substance misuse or domestic and family violence, less than 1% experienced these domains in isolation (Table 3). More than 50% of children experienced five or more disadvantage domains if their parent experienced either justice system contact, substance misuse or domestic and family violence (Table 3).

### Disadvantage co-occurrence within subpopulations

Supplementary Figure 2 to Supplementary Figure 5 show disadvantage profiles among the distinct subgroups of children exposed to parental mental health, substance misuse, domestic family violence and justice system contact. Economic disadvantage was highly prevalent, particularly among children exposed to domestic family violence (97.2%) and justice system contact (89.5%). Children experiencing parental substance misuse were likely to also experience parental mental health disadvantage (95.4%) and vice versa (51.5%). The most commonly co-occurring disadvantages were economic, health, smoking during pregnancy, mental health disadvantage and substance use disadvantage.

### Comparison with area-level measure

Table 4 demonstrates how children experiencing each disadvantage domain were distributed across the area-level socioeconomic disadvantage (IRSAD) quintiles. A high proportion of children whose parents experienced justice system contact (77.3%), domestic and family violence (76.5%), child protection contact (75.9%), low education (75.0%) or economic disadvantage (71.0%) were captured in the two most disadvantaged SEIFA quintiles, though almost a quarter of children who experienced these disadvantages were distributed across the more advantaged IRSAD quintiles. For example, 15.5% of children experiencing economic disadvantage were classified in the two most advantaged IRSAD quintiles.

**Table 4:** Distribution of children experiencing disadvantage from 12 months before birth up to age 5, by Socio-Economic Indexes for Areas (SEIFA) Index of Relative Socioeconomic Advantage and Disadvantage (IRSAD) quintile.

|  | SEIFA IRSAD |  |  |  |  |  |  |  |  |  |
| --- | --- | --- | --- | --- | --- | --- | --- | --- | --- | --- |
|  | 1 |  | 2 |  | 3 |  | 4 |  | 5 |  |
|  | Most disadvantaged |  |  |  |  |  |  |  | Most advantaged |  |
|  | n | % | n | % | n | % | n | % | n | % |
| **Disadvantage domain** |  |  |  |  |  |  |  |  |  |  |
| Economic (n=37,903) | 16,956 | 44.7 | 9,983 | 26.3 | 5,088 | 13.4 | 4,217 | 11.1 | 1,659 | 4.4 |
| Education<sup>(a)</sup> (n=9,113) | 4,521 | 49.6 | 2,317 | 25.4 | 1,189 | 13.1 | 858 | 9.4 | 228 | 2.5 |
| Access to services<sup>(a)</sup> (n=5,432) | 1,174 | 21.6 | 2,820 | 51.9 | 611 | 11.3 | 224 | 4.1 | 603 | 11.1 |
| Mental health (n=18,235) | 7,499 | 41.1 | 4,702 | 25.8 | 2,513 | 13.8 | 2,384 | 13.1 | 1,137 | 6.2 |
| Substance misuse (n=9,780) | 4,512 | 46.2 | 2,426 | 24.8 | 1,236 | 12.7 | 1,119 | 11.5 | 481 | 4.9 |
| Domestic and family violence (n=3,334) | 1,673 | 50.2 | 876 | 26.3 | 392 | 11.8 | 286 | 8.6 | 107 | 3.2 |
| Health (n=29,305) | 10,558 | 36.0 | 7,503 | 25.6 | 4,577 | 15.6 | 4,458 | 15.2 | 2,209 | 7.5 |
| Smoking in pregnancy (n=23,637) | 10,285 | 43.5 | 6,311 | 26.7 | 3,292 | 13.9 | 2,609 | 11.0 | 1,140 | 4.8 |
| Child protection contact<sup>(a)</sup> (n=8,622) | 4,351 | 50.5 | 2,189 | 25.4 | 989 | 11.5 | 799 | 9.3 | 294 | 3.4 |
| Justice System contact (n=2,533) | 1,340 | 52.9 | 617 | 24.4 | 273 | 10.8 | 219 | 8.7 | 84 | 3.3 |
| Death (n=750) | 285 | 38.0 | 192 | 25.6 | 106 | 14.1 | 113 | 15.1 | 54 | 7.2 |
| **Number of domains** |  |  |  |  |  |  |  |  |  |  |
| Not disadvantaged in any domain (n=74,528) | 12,862 | 17.3 | 15,444 | 20.7 | 13,430 | 18.0 | 18,675 | 25.1 | 14,117 | 18.9 |
| 1 domain (n=31,788) | 9,059 | 28.5 | 8,586 | 27.0 | 5,451 | 17.2 | 5,520 | 17.4 | 3,172 | 10.0 |
| 2 domains (n=16,081) | 6,048 | 37.6 | 4,564 | 28.4 | 2,423 | 15.1 | 2,063 | 12.8 | 983 | 6.1 |
| 3 domains (n=8,843) | 3,856 | 43.6 | 2,464 | 27.9 | 1,117 | 12.6 | 1,010 | 11.4 | 396 | 4.5 |
| 4 domains (n=5,158) | 2,456 | 47.6 | 1,392 | 27.0 | 660 | 12.8 | 487 | 9.5 | 163 | 3.2 |
| 5 or more domains (n=6,573) | 3,576 | 54.4 | 1,632 | 24.8 | 703 | 10.7 | 478 | 7.3 | 184 | 2.8 |
| Overall (n=142,971) | 37,857 | 26.5 | 34,082 | 23.8 | 23,784 | 16.6 | 28,233 | 19.8 | 19,015 | 13.3 |

<sup>(a)</sup> Indicators for 'education,' 'access to services' and 'child protection' domains were captured at one point in time and assumed to stay consistent over the duration of six-year study period

Overall, one-sixth of children (17.3%) who were not disadvantaged on any individual-level domain were classified in the most disadvantaged IRSAD quintile. Less than half of children who experienced three (43.6%) or four (47.6%) domains of disadvantage were captured in the most disadvantaged IRSAD quintile. Additionally, only 54.4% of the most multidimensionally disadvantaged children, who experienced disadvantage on five domains or more, were classified in the most disadvantaged IRSAD domain.

## Discussion

In South Australia, almost one in two children (47.9%) were exposed to at least one domain of disadvantage and one in seven (14%) were exposed to three or more disadvantage domains in early life. We found 655 unique combinations of disadvantage domains in the first five years of life which illustrated how disadvantage can manifest and overlap in different ways. Over 50% of children experiencing parental substance misuse, domestic and family violence, or justice system contact also experienced four or more other co-occurring disadvantage domains. The most commonly occurring five-domain combination of disadvantage exposure in early childhood was economic, health, smoking in pregnancy, mental health and substance misuse, which affected 1.3% of children.

Of all disadvantage types, economic disadvantage was the most prevalent. One in four children had parents experiencing economic disadvantage, and 75% of these children experienced an additional disadvantage domain. This is relevant as child welfare literature highlights the triad of parental mental health, substance misuse and domestic and family violence as a common combination responsible for poor child outcomes, but often ignores the influence of socioeconomic disadvantage.^(32)^ The Children’s Commissioner’s Office in England used survey data to estimate that 1.3% of children, up to age five, have an adult in their household experiencing a triad of acute alcohol or drug dependency, severe mental ill-health, and domestic violence in the last year.^(33)^ This is similar to our estimate showing 1.0% of children had parents who had experienced domestic violence, substance misuse and mental health hospitalisation. However, our study found that less than 1% of children who experienced parental substance misuse or domestic and family violence, experienced their disadvantages in isolation, and that more than half of these children experienced five or more disadvantage domains, often co-occurring with economic disadvantage.

Capturing individual-level disadvantage helped us observe how many children may be multidimensionally disadvantaged but live in socioeconomically advantaged areas in South Australia. Approximately half of children exposed to multiple disadvantage domains were not captured in the “most disadvantaged” area-based disadvantage (IRSAD) quintile. Additionally, almost 1 in 6 children exposed to economic disadvantage or smoking during pregnancy were living in areas classified within the two most advantaged IRSAD quintiles. Among children exposed to parental justice system contact or domestic family violence, approximately 12% resided within the two most advantaged IRSAD quintiles. Given that funding for services is often distributed based on disadvantaged priority areas,^(34)^ individual-level views of multidimensional disadvantage can highlight the number of disadvantaged children who may be missing out on services targeted by area. The use of linked administrative data has the potential to support the design and resourcing of services delivered under the principle of proportionate universalism,^(35)^ to ensure they can reach families with complex needs, irrespective of which area they live in.

This paper provides whole-of-population estimates on a range of disadvantage co-occurrences in early life, drawing on individual-level data from systems that respond to different domains of disadvantage. This information can support integration of support systems for children and families in the context of early childhood system reforms.^(22, 36)^ Despite at least 34 countries in the world having some form of integrated provision for children’s services in 2010,^(37)^ evidence gaps and numerous structural challenges raise barriers to successfully achieving an integrated early years system.^(36)^ Data siloes prevent research on the extent of integration required.^(22)^ A review of the international evidence on integrated early years systems highlights that integration needs to be developed from local circumstances and avoid a ‘one size fits all’ approach.^(36)^ As South Australia is currently commencing roll-out of government integrated hubs,^(38)^ this study can inform the context of multidimensional disadvantage of South Australian children.

### Limitations

All constructs of multidimensional disadvantage reflect data availability and decisions as to what constitutes a reasonable indicator of disadvantage. This study presents a conservative estimate of disadvantage as the included indicators often captured acute need for services; for example, parents who had contact with Drug and Alcohol Services reflected a high need for supportive substance misuse services. For the economic indicators, we used proxies for income which likely underestimated the true extent of economic disadvantage. Prevalence of children with a school card or whose parents had low education levels was also likely to be underestimated as data was only available for children who attended government schools and all children who attended non-government schools were counted as not exposed to disadvantage on those indicators.

Given that disadvantaged individuals experience barriers to accessing services, measurement of multidimensional disadvantage may be underestimated as administrative data sources reflect service use. In this study, the ‘access to services’ domain appeared more likely to occur in isolation; however, this may be due to people in remote areas having less service-contact captured in administrative data. Additionally, there are information challenges related to domestic and family violence where numerous barriers prevent disclosure and help-seeking.^(39)^ In this study, indication of domestic and family violence was underestimated as it mainly emerged from public housing data, which also contributed to co-occurrence between economic and domestic violence domains.

Lastly, measurement in this study assumed children were living with their birth parents throughout the first five years of life. Though not the case for all children, there is evidence that caring situations are relatively unchanged for the majority of children; approximately 8.2% of people under 55 years of age experienced separation or divorce in 2021.^(40)^

## Conclusion

Children’s early life exposure to parental disadvantage was diverse and multidimensional. Almost all children with parents experiencing substance misuse, domestic and family violence, or justice system contact experienced additional disadvantages, where disadvantage co-occurrence of economic, health, smoking in pregnancy, mental health and substance misuse was particularly common. These insights, drawn from administrative data on parents’ contact with multiple systems during early childhood, offer an empirical view on the types of system integration and coordination required to comprehensively address families’ needs.

## Data Availability

No data are available for public access. The data underlying this study were provided by several Australian State and Commonwealth government agencies under agreements with the researchers led by author JL, SA NT DataLink as the independent linkage authority and multiple ethics committees. Data are only able to be accessed by researchers who have entered into agreements with the Data Custodians and are approved users by the SA Department for Health and Wellbeing, Human Research Ethics Committee. Data can be accessed through an application and approval process administered by the independent data linkage authority, SA NT DataLink.

## Acknowledgements

We thank the data custodians for providing de-identified data, and SA NT DataLink for undertaking data linkage. This manuscript uses data from the Australia Early Development Census (AEDC). The AEDC is funded by the Australian Government Department of Education. The findings and/or views reported in this study are those of the authors and should not be attributed to any government department.

No competing interests to declare.

