## Supplementary Material for "Early life multidimensional disadvantage of South Australian children: a whole-population linked data study"

### Supplementary Tables

*Supplementary Table 1: Economic disadvantage domain indicators*

| Indicator | Definition | Ages | Data coverage | Data source |
|---|---|---|---|---|
| Family not in labour force at child's birth | If two parents were registered at the child's birth, they were considered unemployed if both parents listed their occupation as student, pensioner, home duties, or unemployed at the time of birth. In lone-parent families, we only considered the occupation listed by the mother. | Birth | Mothers and co-parents registered at child's birth | Perinatal Statistics Collection, South Australian Births Registry |
| Child (or their parent before birth) listed on application in household receiving private rental bond assistance payment | As all public housing information is relative to the head tenant or head applicant, measurement of children's experience of public housing is conditional on them being listed as an occupant of the household.<br>Before the child's birth, disadvantage was determined based on whether parents were listed on an application for rental bond assistance payment. | 12 months before birth up to age 5 | Children (or parents in the year before birth) who had contact with the South Australian Public Housing system | South Australian Public Housing data |
| Child (or their parent before birth) listed either on a waitlist for public housing, or as an occupant in a public housing tenancy | As all public housing information is relative to the head tenant or head applicant, measurement of children's experience of public housing is conditional on them being listed as an occupant of the household.<br>Before the child's birth, disadvantage was determined based on whether parents were listed on a waitlist application or public housing tenancy. | | | |
| Child had school card at school entry | To qualify for a school card, family's gross annual income had to be below a specified limit depending on the number of children in the family and whether the school was government or non-government.<br>Data available on 65,688 children (45.8%). Children not enrolled in government schools or absent during school enrolment census were counted as not having a school card. | Age 5 (school entry) | Children enrolled in government schools in South Australia | Department for Education School Enrolment Census |
| Mother or co-parent had a hospital admission related to unemployment, housing or economic circumstances | Primary or secondary ICD-10-AM diagnoses codes related to unemployment, housing or economic circumstances: Z56.0, Z59.0–Z59.9. | 12 months before birth up to age 5 | Mothers and co-parents admitted to public hospitals in South Australia | Admitted Patient Care data |

*Supplementary Table 2: Education disadvantage domain indicator*

| Indicator | Definition | Ages | Data coverage | Data source |
|---|---|---|---|---|
| Parental highest level of education attained was Year 11 or equivalent or below | This indicator considers if the highest education level of parents was Year 11 or equivalent or below.<br><br>Data available on 65,688 children (45.8%). Children not enrolled in government schools or absent during school enrolment census were counted as having parents with education above Year 11 level.<br><br>Government schools are more likely to have greater number of children whose parents have not completed Year 12. A study using data from the Australian Bureau of Statistics Census of Population and Housing found that 29% of parents with children enrolled in government schools had Year 12 or lower as their highest level of education, compared to 15-20% of parents with children enrolled in non-government schools (Independent Schools Australia, 2023). | Age 5 (school entry) | Children enrolled in government schools in South Australia | Department for Education School Enrolment Census |

*Supplementary Table 3: Access to services disadvantage domain indicator*

| Indicator | Definition | Ages | Data coverage | Data source |
|---|---|---|---|---|
| Mother living in remote or very remote area at child's birth | The Accessibility/Remoteness Index of Australia Plus (ARIA+) divides Australia into five classes of remoteness on the basis of a measure of relative access to services such as health, education or retail (Australian Bureau of Statistics, 2023). The five remoteness classes are: Major Cities, Inner Regional, Outer Regional, Remote and Very Remote.<br><br>While we do not consider living in a remote or very remote area to be a disadvantage in and of itself, it is a proxy for access to services. | Birth<br><br>Assumed to stay consistent from 12 months before birth of child up to age 5 (sensitivity analyses in Supplementary Table 12) | All mothers | Perinatal Statistics Collection, South Australian Births Registry |

| | | |
|---|---|---|
| | | revealed minimal change in remoteness level between birth and age 5. |

*Supplementary Table 4: Mental health disadvantage domain indicators*

| Indicator | Definition | Ages | Data coverage | Data source |
|---|---|---|---|---|
| Mother or co-parent had at least one hospital admission related to mental health | Primary, secondary ICD-10-AM diagnosis or any external cause code related to mental health or self-harm: F10-F19; F20-F29; F30-F39; F40-F48; F50-59; F60-F69; F70-F79; F80-F89; F90-F99; G470; G472; G478; G479; O993; R44; R45-R455; R457-R462; X60-X84. | 12 months before birth up to age 5 | Mothers and co-parents admitted to public hospitals in South Australia | Admitted Patient Care data |
| Mother or co-parent had at least one emergency department presentation related to mental health | Primary ICD-10-AM diagnosis code related to mental health or self-harm: F10-F19; F20-F29; F30-F39; F40-F48; F50-59; F60-F69; F70-F79; F80-F89; F90-F99; G470; G472; G478; G479; O993; R44; R45-R455; R457-R462; X60-X84. | 12 months before birth up to age 5 | Mothers and co-parents who presented to emergency departments in public hospitals in South Australia | Emergency Department Data Collection |

*Supplementary Table 5: Substance misuse disadvantage domain indicators*

| Indicator | Definition | Ages | Data coverage | Data source |
|---|---|---|---|---|
| Mother or co-parent had contact with Drug and Alcohol Services South Australia (DASSA) for their own substance misuse | DASSA is the state-wide South Australian service for people who experience problems with alcohol and other drugs. Parents who have had contact with DASSA have sought or been referred to services relating to substance misuse. | 12 months before birth up to age 5 | Mothers and co-parents who had contact with DASSA for their own substance misuse | Drug and Alcohol Services South Australia |
| Mother or co-parent had at least one hospital admission related to substance misuse | Primary, secondary ICD-10-AM diagnosis or any external cause code related to hospital admission, poisoning, or assault due to substances: F10-F19; T40.0-T40.9; T42.3; T42.4; T42.6; T42.7; T43.3; T43.5; T43.6; T43.8; T43.9; X85-X86; X41; X42; X44; X45; X46; X60-X69; Y10-Y19. | 12 months before birth up to age 5 | Mothers and co-parents admitted to public hospitals in South Australia | Admitted Patient Care data |

| | | | | |
|---|---|---|---|---|
| Mother or co-parent had at least one emergency department presentation related to substance misuse | Primary ICD-10-AM diagnosis code related to hospital admission, poisoning, or assault due to substances: F10-F19; T40.0-T40.9; T42.3; T42.4; T42.6; T42.7; T43.3; T43.5; T43.6; T43.8; T43.9; X85-X86; X41; X42; X44; X45; X46; X60-X69; Y10-Y19. | 12 months before birth up to age 5 | Mothers and co-parents who presented to emergency departments in public hospitals in South Australia | Emergency Department Data Collection |

*Supplementary Table 6: Smoking in pregnancy disadvantage domain indicator*

| Indicator | Definition | Ages | Data coverage | Data source |
|---|---|---|---|---|
| Mother was smoking in pregnancy | Mother was smoking in pregnancy as recorded at the first antenatal appointment (approximately 3 months gestation)<br>Or<br>Mother had hospital admission in six months before child's birth with primary or secondary ICD-10-AM diagnosis or any external cause code related to smoking: P04.2, Z71.6, Z72.0. | Pregnancy to birth | All mothers | Perinatal Statistics Collection, Admitted Patient Care |

*Supplementary Table 7: Domestic and family violence disadvantage domain indicators*

| Indicator | Definition | Ages | Data coverage | Data source |
|---|---|---|---|---|
| Mother or co-parent had at least one hospital admission related to assault, where perpetrator was spouse, domestic partner, parent, other family member, or carer | Primary or secondary ICD-10-AM diagnosis or any external cause codes related to hospital admission, poisoning, or assault due to substances: X9590-X9593; X9920-X9923; X9930-X9933; X9980-X9983; X9990-X9993; Y0230-Y0233; Y0280-Y0283; Y0290-Y0293; Y0320-Y0323; Y0380-Y0383; X8500-X8503; X8600-X8603; X8900-X8903; X9100-X9103; X9300-X9303; X9700-X9703; X9800-X9803; X9900-X9903; Y0000-Y0003; Y0100-Y0103; Y0400-Y0403; Y0500-Y0503; Y0600-Y0603; Y0700-Y0703; Y0800-Y0803; Y0900-Y0903 | 12 months before birth up to age 5 | Mothers and co-parents admitted to public hospitals in South Australia | Admitted Patient Care data |
| Child (or their parent before birth) listed on application in household receiving private rental bond assistance payment, or on waitlist for public housing, or as an occupant in a public housing tenancy where | Children who were listed as an occupant in either:<br>a) A household receiving private rental bond assistance payment where an adult had an indicator of "I am at risk of domestic violence", or<br>b) A household on waitlist for public housing where a reason for applying was due to domestic violence, or<br>c) A household in a public housing tenancy with an intervention order related to domestic violence | 12 months before birth up to age 5 | Children who had contact with the South Australian Public Housing system | South Australian Public Housing data |

| |
|---|
| there was indication of domestic and family violence |

*Supplementary Table 8: Health disadvantage domain indicators*

| **Indicator** | **Definition** | **Ages** | **Data coverage** | **Data source** |
|---|---|---|---|---|
| Mother or co-parent had at least one hospital admission for any cause (except child's birth) that lasted 8 days or more | Hospital admission for 8 days or more within a 12-month period, per parent, was chosen to be indicative of an acute need for health services and acted as a proxy for severe illness or health event. This often accounted for less than 5% of children exposed per each 12-month period between 1 year pre-birth to age 5. More details on the distribution of hospitalisations is provided in Supplementary Table 26 to 27.<br><br>Health disadvantage, measured through hospital admission for 8 days or more, may result in parents who are unavailable or unable to meet their children's needs. Children of parents with a chronic illness may also be at increased risk of negative outcomes including mental and physical health problems, and poorer quality of life (Tossani et al., 2022).<br><br>Hospital admissions where the mother's primary diagnosis was related to the birth or delivery of the child were excluded from this indicator. The following ICD-10-AM codes were used to identify hospitalisation related to birth or delivery: O80-O84; O47-O48; Z37-Z39. | 12 months before birth up to age 5 | Mothers and co-parents admitted to public hospitals in South Australia | Admitted Patient Care data |
| Mother or co-parent experienced 4 or more hospital admissions per year for any cause (except child's birth) | Frequent hospitalisations were classified as 4 or more hospital admissions per parent in a 12-month period, often accounting for less than 2% of children exposed per each 12-month period between 1 year pre-birth to age 5. This was considered to be indicative of an acute need for health services. Data from the Patient Experiences Survey 2016-17 finds that 4.4% of people 15 years or older who had been admitted to hospital had been admitted four or more times (Australian Bureau of Statistics, 2017). | 12 months before birth up to age 5 | Mothers and co-parents admitted to public hospitals in South Australia | Admitted Patient Care data |

| | | | | |
|---|---|---|---|---|
| | More details on the distribution of hospital admissions is provided in Supplementary Table 21 and Supplementary Table 22.<br><br>Hospital admissions where the mother's primary diagnosis was related to the birth or delivery of the child were excluded from this indicator. The following ICD-10-AM codes were used to identify hospitalisation related to birth or delivery: O80-O84; O47-O48; Z37-Z39. | | | |
| Mother or co-parent experienced 4 or more emergency department presentations per year for any cause | Four or more emergency department presentations within a 12-month period was determined to represent an acute need for emergency services, as this often accounted for less than 5% of children exposed per each 12-month period between 1 year pre-birth to age 5. More details on the distribution of emergency department presentations is provided in Supplementary Table 26 and Supplementary Table 27.<br>Frequent emergency department presentations may result in parents who are unavailable or unable to meet their children's needs due to ongoing health constraints. | 12 months before birth up to age 5 | Mothers and co-parents who presented to emergency departments in public hospitals in South Australia | Emergency Department Data Collection |

*Supplementary Table 9: Child protection system contact disadvantage domain indicator*

| Indicator | Definition | Ages | Data coverage | Data source |
|---|---|---|---|---|
| Mother or co-parent had at least one substantiation of maltreatment or experienced out-of-home care placement before child's birth | Instances where young parents experienced child protection contact *after* the child is born were not included. | 12 months before birth | Mothers and co-parents with child protection records in South Australia | Department for Child Protection Data Collection |

*Supplementary Table 10: Justice system contact disadvantage domain indicator*

| Indicator | Definition | Ages | Data coverage | Data source |
|---|---|---|---|---|
| Mother or co-parent experienced imprisonment | Mother or co-parent had contact with the justice system through imprisonment | 12 months before birth up to age 5 | Mothers and co-parents with adult criminal history records in South Australia | Corrections Data |

*Supplementary Table 11: Parental death disadvantage domain indicator*

| Indicator | Definition | Ages | Data coverage | Data source |
|---|---|---|---|---|
| Mother or co-parent died | Mother or co-parent listed in deaths registry | 12 months before birth up to age 5 | Mothers and co-parents with death registered in South Australia | South Australian Deaths Registry |

NB: Each indicator was measured from 12 months before the child's birth up to age 5, with the exception of five indicators (jobless family, parental education, remoteness of area of residence, smoking in pregnancy, and parents' history of contact with the child protection system) that were only recorded at the specific time points listed in the tables above.

*Supplementary Table 12: Sensitivity Analysis on living in a remote or very remote area at birth and at age of school entry (n=35,830)*

| | **Child not living in a remote or very remote area at school entry** | | **Child living in a remote or very remote area at school entry** | | |
|---|---|---|---|---|---|
| | n | % of total children | n | % of total children | Total |
| **Mother not living in a remote or very remote area at child's birth** | 34,027 | 98.8 | 406 | 1.2 | 34,433 |
| **Mother living in a remote or very remote area at child's birth** | 359 | 25.7 | 1038 | 74.3 | 1,397 |
| Total | 34,386 | 96.0 | 1444 | 4.0 | 35,830 |

The *access to services* domain included whether the child's mother was living in a remote or very remote area at child's birth. This was compared to whether the child was living in a remote or very remote area at age of school entry in a subset of the population, as collected in the Australian Early Development Census in 2009, 2010, 2012, 2015 and 2018.

Overall, a total of 2.1% of all 35,830 children would be misclassified on living in a remote or very remote area throughout childhood if only the measure of remoteness at birth is used.

*Supplementary Table 13: Persistent disadvantage – children, n (%), number of years in which domain-specific disadvantage was observed*

| Parental disadvantage domain[a] | 1 year | | 2 years | | 3 years | | 4 years | | 5 years | | 6 years | |
|---|---|---|---|---|---|---|---|---|---|---|---|---|
| | n | % | n | % | n | % | n | % | n | % | n | % |
| Economic (n=37,924) | 14,778 | 39.0 | 7,818 | 20.6 | 5,400 | 14.2 | 3,857 | 10.2 | 3,046 | 8.0 | 3,025 | 8.0 |
| Mental Health (n=18,248) | 12,518 | 68.6 | 3,462 | 19.0 | 1,317 | 7.2 | 564 | 3.1 | 285 | 1.6 | 102 | 0.6 |
| Substance misuse (n=9,780) | 6,820 | 69.7 | 1,740 | 17.8 | 710 | 7.3 | 305 | 3.1 | 154 | 1.6 | 51 | 0.5 |
| Domestic and family violence (n=3,335) | 2,209 | 66.2 | 725 | 21.7 | 280 | 8.4 | 92 | 2.8 | n.d | n.d | n.d | n.d |
| Health (n=29,327) | 21,604 | 73.7 | 5,091 | 17.4 | 1,612 | 5.5 | 602 | 2.1 | 282 | 1.0 | 136 | 0.5 |
| Justice System contact (n=2,534) | 1,286 | 50.8 | 529 | 20.9 | 304 | 12.0 | 192 | 7.6 | 122 | 4.8 | 101 | 4.0 |

n.d. not disclosed due to identification risk of low number of individuals

[a] Persistence was not calculated for domains where information was not measured over time (education, access to services, smoking in pregnancy, and parental child protection system contact)

Supplementary Table 13 demonstrates that the majority of children exposed to parental health disadvantage (73.7%), substance misuse (69.7%), or mental health disadvantage (68.6%) were likely to only be exposed to the disadvantage in a single year. Conversely, 61% of children exposed to economic disadvantage experienced recurrence of this disadvantage over time, with 8.0% exposed every year from 12 months before birth to age 5.

However, persistence of disadvantage should be interpreted with caution as measurement primarily relied on parents' interactions with support systems. For example, parents may have an ongoing mental health condition but only present to hospital in one year; absence of system contact does not necessarily mean there is an absence of disadvantage in that year.

*Supplementary Table 14: Economic, education, and access to services disadvantage – children, n (%), experiencing indicators from 12 months before birth up to age 5*

| | Birth (n=143,378) | Age 1 (143,180) | Age 2 (n=143,128) | Age 3 (n=143,108) | Age 4 (n=143,098) | Age 5 (n=143,083) | Overall from 12 months before birth to age 5 (n=143,083) |
|---|---|---|---|---|---|---|---|
| **Economic** | | | | | | | |
| Child listed on application in household receiving private rental bond assistance | - | 8,475 (5.9) | 8,410 (5.9) | 7,813 (5.5) | 7,516 (5.3) | 7,176 (5.0) | 22,285 (15.6) |
| Mother listed on application in household receiving private rental bond assistance | 5,118 (3.6) | - | - | - | - | - | 5,096 (3.6) |
| Co-parent listed on application in household receiving private rental bond assistance | 2,039 (1.4) | - | - | - | - | - | 2,031 (1.4) |
| Child had school card at age of school entry | - | - | - | - | - | 18,111 (12.7) | 18,111 (12.7) |
| Child in public housing or registered on public housing waitlist | - | 7,559 (5.3) | 7,180 (5.0) | 6,967 (4.9) | 6,683 (4.7) | 6,529 (4.6) | 13,997 (9.8) |
| Mother in public housing or registered on public housing waitlist | 7,413 (5.2) | - | - | - | - | - | 7,373 (5.2) |
| Co-parent in public housing or registered on public housing waitlist | 3,539 (2.5) | - | - | - | - | - | 3,524 (2.5) |
| Family had hospital admission related to problems with income or housing | 262 (0.2) | 257 (0.2) | 200 (0.1) | 217 (0.2) | 206 (0.1) | 259 (0.2) | 1,254 (0.9) |
| Child in jobless family | 11,712 (8.2) | - | - | - | - | - | 11,647 (8.1) |
| Family had hospital admission related to unemployment | 18 (0.0) | 23 (0.0) | 37 (0.0) | 35 (0.0) | 32 (0.0) | 36 (0.0) | 172 (0.1) |
| **Education** | | | | | | | |
| Parents' highest level of education attained was Year 11 or equivalent or below | - | - | - | - | - | 9,118 (6.4) | 9,118 (6.4) |
| **Access to services** | | | | | | | |
| Mother living in geographically remote or very remote area | 5,475 (3.8) | - | - | - | - | - | 5,462 (3.8) |

*Supplementary Table 15: Mental health disadvantage – children, n (%), whose mothers experienced indicators from 12 months before birth up to age 5*

| | **Birth (n=143,378)** | **Age 1 (143,180)** | **Age 2 (n=143,128)** | **Age 3 (n=143,108)** | **Age 4 (n=143,098)** | **Age 5 (n=143,083)** | **Overall from 12 months before birth to age 5 (n=143,083)** |
|---|---|---|---|---|---|---|---|
| Mother had hospital admission related to mental health | 3,761 (2.6) | 4,053 (2.8) | 1,814 (1.3) | 1,881 (1.3) | 1,776 (1.2) | 1,778 (1.2) | 10,918 (7.6) |
| Mother had emergency department presentation related to mental health | 940 (0.7) | 1,060 (0.7) | 1,030 (0.7) | 1,103 (0.8) | 1,244 (0.9) | 1,378 (1.0) | 4,993 (3.5) |

*Supplementary Table 16: Mental health disadvantage – children, n (%), whose co-parent experienced indicators from 12 months before birth up to age 5*

| | **Birth (n=139,534)** | **Age 1 (139,347)** | **Age 2 (n=139,297)** | **Age 3 (n=139,277)** | **Age 4 (n= 139,268)** | **Age 5 (n=139,255)** | **Overall from 12 months before birth to age 5 (n= 139,255)** |
|---|---|---|---|---|---|---|---|
| Co-parent had hospital admission related to mental health | 875 (0.6) | 1,118 (0.8) | 1,244 (0.9) | 1,294 (0.9) | 1,262 (0.9) | 1,342 (1.0) | 5,211 (3.7) |
| Co-parent had emergency department presentation related to mental health | 673 (0.5) | 902 (0.7) | 979 (0.7) | 1,102 (0.8) | 1,129 (0.8) | 1,175 (0.8) | 4,439 (3.2) |

*Supplementary Table 17: Substance misuse disadvantage – children, n (%), whose mothers experienced indicators from 12 months before birth up to age 5*

| | Birth (n=143,378) | Age 1 (143,180) | Age 2 (n=143,128) | Age 3 (n=143,108) | Age 4 (n=143,098) | Age 5 (n=143,083) | Overall from 12 months before birth to age 5 (n=143,083) |
|---|---|---|---|---|---|---|---|
| Mother had contact with DASSA services | 88 (0.1) | 73 (0.1) | 66 (0.1) | 72 (0.1) | 81 (0.1) | 80 (0.1) | 313 (0.2) |
| Mother had hospital admission related to substance misuse | 1,687 (1.2) | 1,894 (1.3) | 949 (0.7) | 912 (0.6) | 951 (0.7) | 959 (0.7) | 5,251 (3.7) |
| Mother had ED presentation related to substance misuse | 180 (0.1) | 223 (0.2) | 274 (0.2) | 328 (0.2) | 352 (0.3) | 398 (0.3) | 1,394 (1.0) |

*Supplementary Table 18: Substance misuse disadvantage – children, n (%), whose co-parent experienced indicators from 12 months before birth up to age 5*

| | Birth (n=139,534) | Age 1 (139,347) | Age 2 (n= 139,297) | Age 3 (n=139,277) | Age 4 (n= 139,268) | Age 5 (n=139,255) | Overall from 12 months before birth to age 5 (n= 139,255) |
|---|---|---|---|---|---|---|---|
| Co-parent had contact with DASSA services | 87 (0.1) | 111 (0.1) | 102 (0.1) | 101 (0.1) | 79 (0.1) | 81 (0.1) | 398 (0.3) |
| Co-parent had hospital admission related to substance misuse | 682 (0.5) | 839 (0.6) | 931 (0.7) | 941 (0.7) | 927 (0.7) | 970 (0.7) | 3,898 (2.8) |
| Co-parent had ED presentation related to substance misuse | 229 (0.2) | 280 (0.2) | 335 (0.2) | 376 (0.3) | 356 (0.3) | 408 (0.3) | 1,619 (1.2) |

*Supplementary Table 19: Domestic and family violence disadvantage – children, n (%), who experienced indicators from 12 months before birth up to age 5*

| | Birth (n=143,378) | Age 1 (143,180) | Age 2 (n=143,128) | Age 3 (n=143,108) | Age 4 (n=143,098) | Age 5 (n=143,083) | Overall from 12 months before birth to age 5 (n=143,083) |
|---|---|---|---|---|---|---|---|
| Child listed on application in household receiving private rental bond assistance payment, or on waitlist for public housing, or as an occupant in a public housing tenancy where there was indication of domestic and family violence | - | 690 (0.5) | 753 (0.5) | 754 (0.5) | 717 (0.5) | 706 (0.5) | 2,398 (1.7) |
| Mother listed on application in household receiving private rental bond assistance payment, or on waitlist for public housing, or as an occupant in a public housing tenancy where there was indication of domestic and family violence | 507 (0.4) | - | - | - | - | - | 504 (0.4) |
| Mother had hospital admission related to domestic and family violence | 178 (0.1) | 134 (0.1) | 129 (0.1) | 126 (0.1) | 119 (0.1) | 134 (0.1) | 713 (0.5) |

*Supplementary Table 20: Domestic and family violence disadvantage – children, n (%), whose co-parent experienced indicators from 12 months before birth up to age 5*

| | Birth (n=139,534) | Age 1 (139,347) | Age 2 (n=139,297) | Age 3 (n=139,277) | Age 4 (n=139,268) | Age 5 (n=139,255) | Overall from 12 months before birth to age 5 (n= 139,255) |
|---|---|---|---|---|---|---|---|
| Co-parent listed on application in household receiving private rental bond assistance payment, or on waitlist for public housing, or as an occupant in a public housing tenancy where there was indication of domestic and family violence | 33 (0.0) | - | - | - | - | - | 32 (0.0) |
| Co-parent had hospital admission related to domestic and family violence | 30 (0.0) | 26 (0.0) | 46 (0.0) | 26 (0.0) | 22 (0.0) | 21 (0.0) | 160 (0.1) |

*Supplementary Table 21: All cause health disadvantage – children, n (%) unless specified otherwise, whose mothers experienced indicators from 12 months before birth up to age 5*

| | | Birth (n=143,378) | Age 1 (143,180) | Age 2 (n=143,128) | Age 3 (n=143,108) | Age 4 (n=143,098) | Age 5 (n=143,083) | Overall from 12 months before birth to age 5 (n=143,083) |
|---|---|---|---|---|---|---|---|---|
| Mother had any hospital admission within 12-month period | | 47,869 (33.4) | 50,094 (35.0) | 27,198 (19.0) | 26,423 (18.5) | 23,898 (16.7) | 21,534 (15.1) | 111,843 (78.2) |
| Number of hospitalisations within 12-month period | None | 95,509 (66.6) | 93,086 (65.0) | 115,930 (81.0) | 114,316 (79.9) | 119,200 (83.3) | 121,549 (85.0) | 42,294 (29.6) |
| | 1 | 32,526 (22.7) | 40,852 (28.5) | 20,119 (14.1) | 21,446 (15.0) | 17,537 (12.3) | 15,815 (11.1) | 90,300 (63.1) |
| | 2 | 9,616 (6.7) | 6,680 (4.7) | 4,708 (3.3) | 4,825 (3.4) | 4,181 (2.9) | 3,711 (2.6) | 28,447 (19.9) |
| | 3 | 3,300 (2.3) | 1,598 (1.1) | 1,441 (1.0) | 1,500 (1.1) | 1,211 (0.9) | 1,085 (0.8) | 9,218 (6.4) |
| | 4 or more | 2,427 (1.7) | 964 (0.7) | 930 (0.7) | 1,021 (0.7) | 969 (0.7) | 923 (0.7) | 5,978 (4.2) |
| Length of stay (days) of mother's hospital admissions within 12-month period | Mean (SD) | 4.9 (6.6) | 3.79 (5.3) | 3.5 (6.2) | 3.5 (6.7) | 3.5 (6.9) | 3.6 (7.5) | 8.1 (12.3) |
| | Min - max | 1-258 | 1-227 | 1-254 | 1-334 | 1-273 | 1-372 | 1-699 |
| | IQR | 1-6 | 2-4 | 1-4 | 1-4 | 1-4 | 1-4 | 3-9 |
| | Median | 3 | 3 | 2 | 2 | 2 | 2 | 6 |
| Length of stay for mother's hospital admissions within 12-month period | None | 95,509 (66.6) | 93,086 (65.0) | 115,930 (81.0) | 114,316 (79.9) | 119,200 (83.3) | 121,549 (85.0) | 42,294 (29.6) |
| | 1 day | 12,828 (9.0) | 11,123 (7.8) | 10,429 (7.3) | 10,418 (7.3) | 9,627 (6.7) | 9,245 (6.5) | 48,900 (34.2) |
| | 2-3 days | 11,629 (8.11) | 18,890 (13.2) | 8,962 (6.3) | 9,984 (7.0) | 7,728 (5.4) | 6,709 (4.7) | 50,251 (35.1) |
| | 4-7 days | 16,505 (11.5) | 17,084 (11.9) | 5,912 (4.1) | 6,402 (4.5) | 4,729 (3.3) | 3,886 (2.7) | 44,021 (30.8) |
| | 8-14 days | 4,846 (3.4) | 2,098 (1.5) | 1,211 (0.9) | 1,253 (0.9) | 1,152 (0.8) | 1,021 (0.7) | 10,479 (7.3) |
| | >2 weeks | 2,061 (1.4) | 899 (0.6) | 684 (0.5) | 735 (0.5) | 662 (0.5) | 673 (0.5) | 4,796 (3.4) |

| | | | | | | | | |
|---|---|---|---|---|---|---|---|---|
| Mother had any ED presentation within 12-month period | Yes | 44,024 (30.7) | 32,446 (22.7) | 21,830 (15.3) | 22,171 (15.5) | 21,360 (15.0) | 21,249 (14.9) | 81,704 (57.1) |
| Mother's number of ED presentations within 12-month period | None | 99,354 (69.3) | 110,734 (77.3) | 121,298 (84.8) | 120,937 (84.5) | 121,738 (85.1) | 121,834 (85.2) | 61,379 (42.9) |
| | 1 | 20,886 (14.6) | 22,264 (15.6) | 13,504 (9.4) | 13,780 (9.6) | 13,513 (9.4) | 13,673 (9.6) | 66,413 (46.4) |
| | 2 | 9,978 (7.0) | 6,041 (4.2) | 4,382 (3.1) | 4,455 (3.1) | 4,238 (3.0) | 4,131 (2.9) | 27,257 (19.1) |
| | 3 | 5,329 (3.7) | 2,060 (1.4) | 1,852 (1.3) | 1,784 (1.3) | 1,709 (1.1) | 1,599 (1.1) | 12,731 (8.9) |
| | 4 or more | 7,831 (5.5) | 2,081 (1.5) | 2,092 (1.5) | 2,152 (1.5) | 1,900 (1.3) | 1,846 (1.3) | 13,517 (9.5) |

*Supplementary Table 22: All cause health disadvantage – children, n (%) unless specified otherwise, whose co-parents experienced indicators from 12 months before birth up to age 5*

| | | Birth (n=139,534) | Age 1 (139,347) | Age 2 (n=139,297) | Age 3 (n=139,277) | Age 4 (n= 139,268) | Age 5 (n=139,255) | Overall from 12 months before birth to age 5 (n= 139,255) |
|---|---|---|---|---|---|---|---|---|
| Co-parent had any hospital admission within 12-month period | | 7,216 (5.2) | 9,325 (6.7) | 9,056 (6.5) | 9,230 (6.6) | 9,186 (6.6) | 9,262 (6.6) | 37,919 (27.1) |
| Number of hospitalisations within 12-month period | None | 132,330 (94.8) | 130,068 (93.3) | 130,282 (93.5) | 130,093 (93.5) | 130,115 (93.4) | 130,029 (93.4) | 101,501 (72.9) |
| | 1 | 5,696 (4.1) | 7,453 (5.4) | 7,020 (5.0) | 7,163 (5.0) | 7,094 (5.1) | 7,107 (5.1) | 32,928 (23.7) |
| | 2 | 1,059 (0.8) | 1,230 (0.9) | 1,329 (1.0) | 1,309 (1.0) | 1,317 (0.9) | 1,377 (1.0) | 6,842 (4.9) |
| | 3 | 262 (0.2) | 338 (0.2) | 379 (0.3) | 353 (0.3) | 399 (0.3) | 398 (0.3) | 1,983 (1.4) |
| | 4 or more | 187 (0.1) | 258 (0.2) | 287 (0.2) | 359 (0.2) | 343 (0.3) | 344 (0.3) | 1,394 (1.0) |
| Length of stay (days) of co-parent's hospital admissions within 12-month period | Mean (SD) | 3.1 (7.6) | 3.4 (11.0) | 3.7 (10.1) | 3.8 (11.5) | 3.9 (11.2) | 3.9 (10.4) | 5.1 (17.3) |
| | Min - max | 1-186 | 1-387 | 1-248 | 1-375 | 1-382 | 1-265 | 1-908 |
| | IQR | 1-3 | 1-2 | 1-3 | 1-3 | 1-3 | 1-3 | 1-4 |
| | Median | 1 | 1 | 1 | 1 | 1 | 1 | 2 |
| Length of stay for co-parent's hospital admissions within 12-month period | None | 132,594 (94.8) | 130,485 (93.3) | 130,754 (93.5) | 130,580 (93.4) | 130,624 (93.4) | 130,548 (93.4) | 101,501 (72.9) |
| | 1 day | 4,151 (3.0) | 5,771 (4.1) | 5,336 (3.8) | 5,376 (3.9) | 5,401 (3.9) | 5,319 (3.8) | 26,222 (18.8) |
| | 2-3 days | 1,721 (1.2) | 1,928 (1.4) | 1,940 (1.4) | 2,021 (1.5) | 1,962 (1.4) | 1,997 (1.4) | 10,330 (7.4) |
| | 4-7 days | 799 (0.6) | 918 (0.7) | 962 (0.7) | 988 (0.7) | 995 (0.7) | 1,005 (0.7) | 5,107 (3.7) |
| | 8-14 days | 341 (0.2) | 402 (0.3) | 444 (0.3) | 434 (0.3) | 422 (0.3) | 499 (0.4) | 2,346 (1.7) |
| | >2 weeks | 204 (0.2) | 306 (0.2) | 374 (0.3) | 411 (0.3) | 406 (0.3) | 442 (0.3) | 1,667 (1.2) |

| | | | | | | | | |
|---|---|---|---|---|---|---|---|---|
| Co-parent had any ED presentation within 12-month period | Yes | 15,062 (10.8) | 15,225 (10.9) | 16,094 (11.6) | 16,858 (12.1) | 17,055 (12.3) | 17,852 (12.8) | 57,128 (41.0) |
| Co-parent's number of ED presentations within 12-month period | None | 124,472 (89.2) | 124,122 (89.1) | 123,203 (88.5) | 122,419 (87.9) | 122,213 (87.8) | 121,403 (87.2) | 82,127 (59.0) |
| | 1 | 11,001 (7.9) | 10,903 (7.8) | 11,401 (8.2) | 11,916 (8.6) | 12,076 (8.7) | 12,576 (9.0) | 49,455 (35.1) |
| | 2 | 2,617 (1.9) | 2,768 (2.0) | 2,892 (2.1) | 3,116 (2.2) | 3,083 (2.2) | 3,290 (2.4) | 14,933 (10.7) |
| | 3 | 817 (0.6) | 888 (0.6) | 991 (0.7) | 997 (0.7) | 1,032 (0.7) | 1,080 (0.8) | 5,211 (3.7) |
| | 4 or more | 627 (0.5) | 666 (0.5) | 810 (0.6) | 829 (0.6) | 864 (0.6) | 906 (0.7) | 3,565 (2.6) |

*Supplementary Table 23: Smoking during pregnancy – children, n (%), whose mothers experienced indicators from 12 months before birth up to age 5*

| | **Birth (n=143,378)** | **Age 1 (143,180)** | **Age 2 (n=143,128)** | **Age 3 (n=143,108)** | **Age 4 (n=143,098)** | **Age 5 (n=143,083)** | **Overall from 12 months before birth to age 5 (n=143,083)** |
|---|---|---|---|---|---|---|---|
| Mother was smoking in pregnancy at first antenatal appointment | 23,760 (16.6) | - | - | - | - | - | 23,652 (16.5) |
| Mother had hospital admission in six months before child's birth with ICD-10-AM diagnosis or external cause code related to smoking | 16,547 (11.5) | - | - | - | - | - | 16,468 (11.5) |

*Supplementary Table 24: Child protection contact – children, n (%), whose mothers experienced indicators from 12 months before birth up to age 5*

| | **Birth (n=143,378)** | **Age 1 (143,180)** | **Age 2 (n=143,128)** | **Age 3 (n=143,108)** | **Age 4 (n=143,098)** | **Age 5 (n=143,083)** | **Overall from 12 months before birth to age 5 (n=143,083)** |
|---|---|---|---|---|---|---|---|
| Mother ever had substantiation of maltreatment or out-of-home care placement before child's birth | 6,360 (4.4) | - | - | - | - | - | 6,335 (4.4) |

*Supplementary Table 25: Child protection contact – children, n (%), whose co-parents experienced indicators from 12 months before birth up to age 5*

| | **Birth (n=139,534)** | **Age 1 (139,347)** | **Age 2 (n=139,297)** | **Age 3 (n=139,277)** | **Age 4 (n=139,268)** | **Age 5 (n=139,255)** | **Overall from 12 months before birth to age 5 (n= 139,255)** |
|---|---|---|---|---|---|---|---|
| Co-parent ever had substantiation of maltreatment or out-of-home care placement before child's birth | 2,922 (2.0) | - | - | - | - | - | 2,908 (2.0) |

*Supplementary Table 26: Justice system contact – children, n (%), whose mothers experienced indicators from 12 months before birth up to age 5*

| | Birth (n=143,378) | Age 1 (143,180) | Age 2 (n=143,128) | Age 3 (n=143,108) | Age 4 (n=143,098) | Age 5 (n=143,083) | Overall from 12 months before birth to age 5 (n=143,083) |
|---|---|---|---|---|---|---|---|
| Mother experienced imprisonment | 130<br>(0.1) | 123<br>(0.1) | 173<br>(0.1) | 119<br>(0.1) | 239<br>(0.2) | 251<br>(0.2) | 578<br>(0.4) |

*Supplementary Table 27: Justice system contact – children, n (%), whose co-parents experienced indicators from 12 months before birth up to age 5*

| | Birth (n=139,534) | Age 1 (139,347) | Age 2 (n=139,297) | Age 3 (n=139,277) | Age 4 (n=139,268) | Age 5 (n=139,255) | Overall from 12 months before birth to age 5 (n= 139,255) |
|---|---|---|---|---|---|---|---|
| Co-parent experienced imprisonment | 507<br>(0.4) | 605<br>(0.4) | 681<br>(0.5) | 714<br>(0.5) | 816<br>(0.6) | 898<br>(0.6) | 2,049<br>(1.5) |

*Supplementary Table 28: Children, n (%), who experienced death of mother from 12 months before birth up to age 5*

| | Birth (n=143,378) | Age 1 (143,180) | Age 2 (n=143,128) | Age 3 (n=143,108) | Age 4 (n=143,098) | Age 5 (n=143,083) | Overall from 12 months before birth to age 5 (n=143,083) |
|---|---|---|---|---|---|---|---|
| Mother death | 0 (0.0) | 22 (0.0) | 33 (0.0) | 43 (0.0) | 48 (0.0) | 63 (0.0) | 209 (0.2) |

*Supplementary Table 29: Children, n (%), who experienced death of co-parent from 12 months before birth up to age 5*

| | Birth (n=139,534) | Age 1 (139,347) | Age 2 (n= 139,297) | Age 3 (n=139,277) | Age 4 (n= 139,268) | Age 5 (n=139,255) | Overall from 12 months before birth to age 5 (n= 139,255) |
|---|---|---|---|---|---|---|---|
| Co-parent death | 49 (0.0) | 89 (0.1) | 106 (0.1) | 98 (0.1) | 96 (0.1) | 109 (0.1) | 498 (0.4) |

Mothers had consistently higher proportions of hospital admission or emergency department (ED) presentations relating to mental health, domestic or family violence, or all cause health disadvantage, than co-parents (Supplementary Table 15 to Supplementary Table 16, Supplementary Table 19 to Supplementary Table 22). Parental death, however, was more prevalent among co-parents than mothers (Supplementary Table 28 to Supplementary Table 29).

*Supplementary Table 30: Children, n (%), who ever experienced domain-specific disadvantage, by age*

| Parental disadvantage domain | 12 months before birth (n=143,378) | | Age 1 (143,180) | | Age 2 (n=143,128) | | Age 3 (n=143,108) | | Age 4 (n=143,098) | | Age 5 (n=143,083) | |
|---|---|---|---|---|---|---|---|---|---|---|---|---|
| | n | % | n | % | n | % | n | % | n | % | n | % |
| Economic | 18,393 | 12.8 | 13,464 | 9.4 | 13,537 | 9.5 | 13,043 | 9.1 | 12,665 | 8.9 | 29,430 | 17.1 |
| Education[(a)] | 9,118 | 6.4 | 9,118 | 6.4 | 9,118 | 6.4 | 9,118 | 6.4 | 9,118 | 6.4 | 9,118 | 6.4 |
| Access to services[(a)] | 5,475 | 3.8 | 5,466 | 3.8 | 5,464 | 3.8 | 5,463 | 3.8 | 5,463 | 3.8 | 5,462 | 3.8 |
| Mental health | 5,350 | 3.7 | 5,919 | 4.1 | 3,942 | 2.8 | 4,138 | 2.9 | 4,129 | 2.9 | 4,250 | 3.0 |
| Substance misuse | 2,617 | 1.8 | 3,022 | 2.1 | 2,217 | 1.6 | 2,262 | 1.6 | 2,276 | 1.6 | 2,361 | 1.7 |
| Domestic and family violence | 698 | 0.5 | 836 | 0.6 | 906 | 0.6 | 891 | 0.6 | 847 | 0.6 | 847 | 0.6 |
| Health | 14,773 | 10.3 | 6,059 | 4.2 | 5,189 | 3.6 | 5,443 | 3.8 | 4,986 | 3.5 | 4,876 | 3.4 |
| Smoking in pregnancy[(b)] | 23,760 | 16.6 | | | | | | | | | | |
| Child protection contact[(a)] | 8,665 | 6.0 | 8,638 | 6.0 | 8,634 | 6.0 | 8,633 | 6.0 | 8,630 | 6.0 | 8,630 | 6.0 |
| Justice system contact | 625 | 0.4 | 717 | 0.5 | 838 | 0.6 | 903 | 0.6 | 1,030 | 0.7 | 1,129 | 0.8 |
| Death | 49 | 0.0 | 110 | 0.1 | 139 | 0.1 | 141 | 0.1 | 144 | 0.1 | 171 | 0.1 |

[(a)] Indicators for ‘education,’ ‘access to services’ and ‘child protection’ domains were captured at one point in time and assumed to stay consistent over the duration of six-year study period

[(b)] Smoking during pregnancy was only included in the age-specific analysis before birth and not at ages 1 to 5

Supplementary Table 30 shows the age-specific prevalence of each disadvantage domain. Parental health disadvantage was most common before the child’s birth (10.3%) and less prevalent in later years (3.4% – 4.2%). Exposure to parental mental health disadvantage affected 12.8% over the six-year period (Table 1) but only 2.8% to 4.1% experienced this disadvantage domain in any specific year of age.

*Supplementary Figure 1: Population Flow Chart*

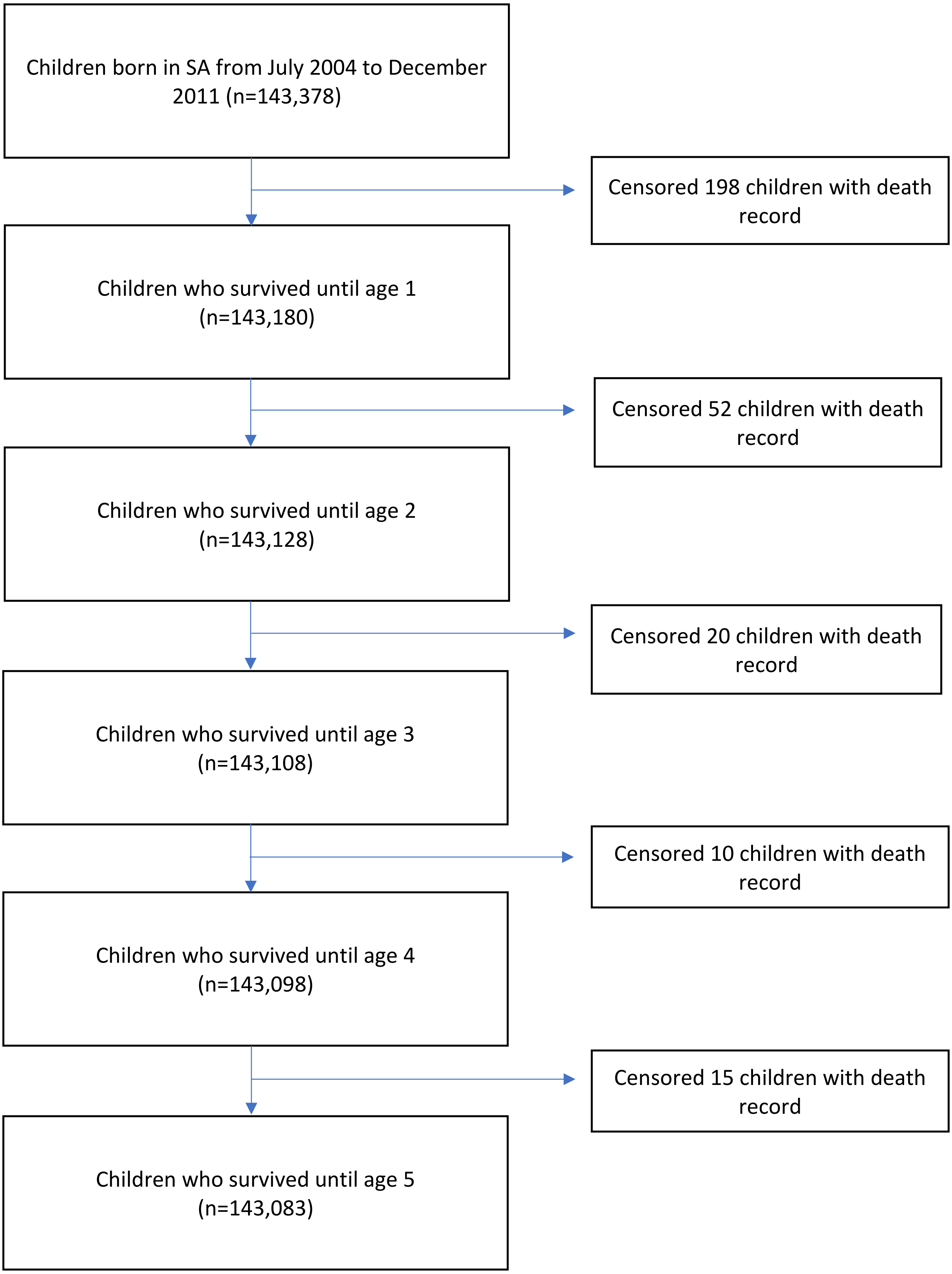

*Supplementary Figure 2: Prevalence of each disadvantage domain, and the 20 most prevalent combinations of disadvantage domains (out of total 286 combinations) for children who experienced Parental Substance Misuse disadvantage from 12 months before birth to age 5 (n=9,780)*

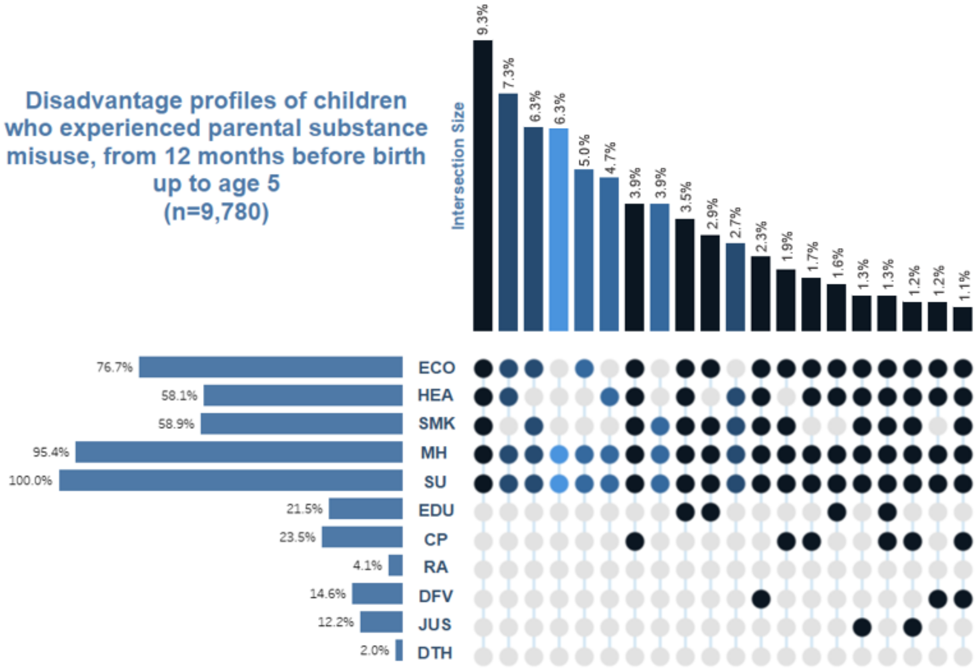

Domain abbreviations: ECO = economic disadvantage, HEA = health disadvantage, SMK = smoking during pregnancy, MH = mental health disadvantage, SU = substance misuse disadvantage, EDU = education disadvantage, CP = child protection disadvantage, RA = access to services disadvantage, DFV = domestic family violence, JUS = justice system contact; DTH = death of parent

Supplementary Figure 2 shows that, among children who experienced parental substance misuse, mental health was the most prevalent co-occurring disadvantage (95.4%), followed by economic disadvantage (76.7%). The most prevalent combination of disadvantage experienced (9.3%) was a five-domain combination of economic, health, smoking during pregnancy, and mental health disadvantage.

*Supplementary Figure 3: Prevalence of each disadvantage domain, and the 20 most prevalent combinations of disadvantage domains (out of total 360 combinations) for children who experienced Parental Mental Health disadvantage from 12 months before birth to age 5 (n=18.248)*

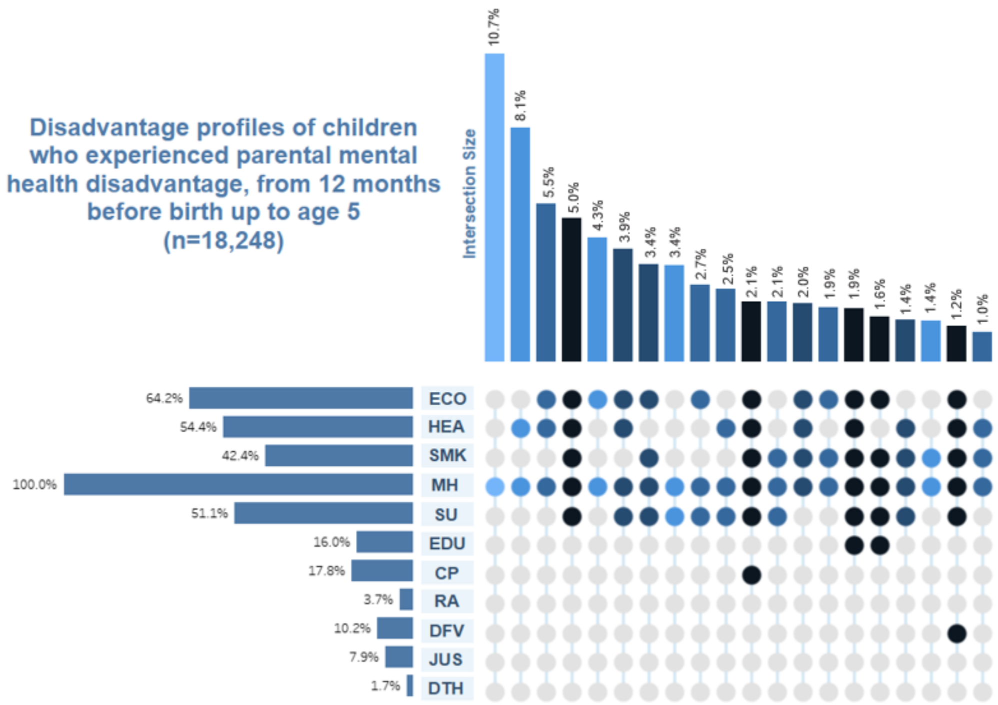

Domain abbreviations: ECO = economic disadvantage, HEA = health disadvantage, SMK = smoking during pregnancy, MH = mental health disadvantage, SU = substance misuse disadvantage, EDU = education disadvantage, CP = child protection disadvantage, RA = access to services disadvantage, DFV = domestic family violence, JUS = justice system contact; DTH = death of parent

Supplementary Figure 3 shows that the domains most likely to co-occur with mental health disadvantage were economic disadvantage (64.2%), health disadvantage (54.4%) and substance misuse (51.1%).

*Supplementary Figure 4: Prevalence of each disadvantage domain, and the 20 most prevalent combinations of disadvantage domains (out of total 210 combinations) for children who experienced Domestic and Family Violence disadvantage from 12 months before birth to age 5 (n=3,335)*

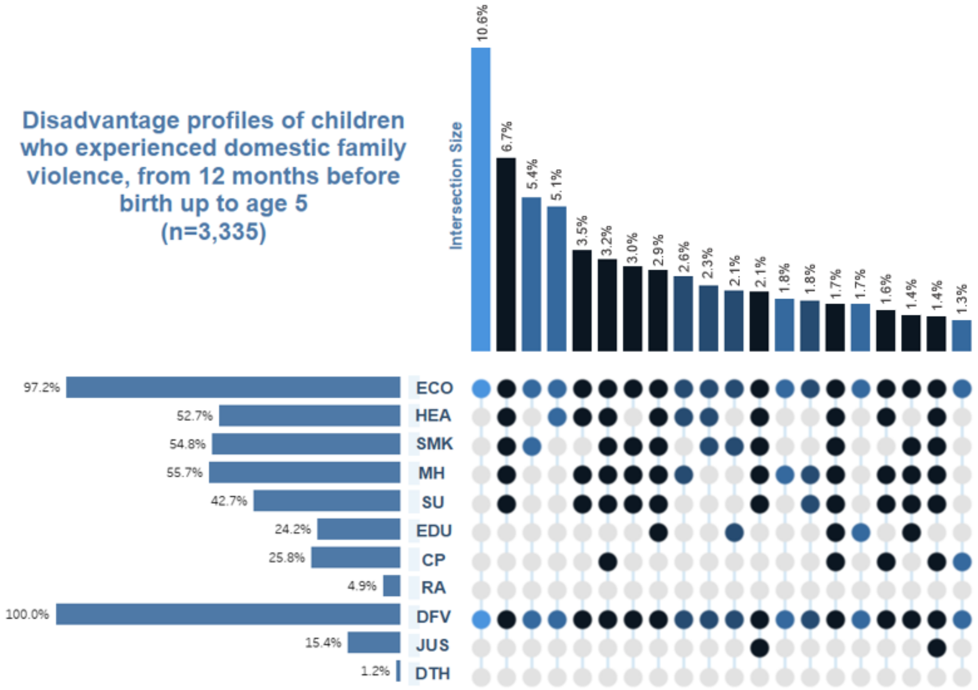

Domain abbreviations: ECO = economic disadvantage, HEA = health disadvantage, SMK = smoking during pregnancy, MH = mental health disadvantage, SU = substance misuse disadvantage, EDU = education disadvantage, CP = child protection disadvantage, RA = access to services disadvantage, DFV = domestic family violence, JUS = justice system contact; DTH = death of parent

Supplementary Figure 4 demonstrates that, among children who experienced domestic and family violence, the majority (97.2%) experienced economic disadvantage, with the two-domain co-occurrence of domestic and family violence with economic disadvantage being the most prevalent combination (10.6%). The second most prevalent combination among children who experienced domestic family violence (6.7%) was a 6-domain experience of economic, health, smoking in pregnancy, mental health and parental substance misuse.

*Supplementary Figure 5: Prevalence of each disadvantage domain, and the 20 most prevalent combinations of disadvantage domains (out of total 228 combinations) for children who experienced Parental Justice System disadvantage from 12 months before birth to age 5 (n=2,534)*

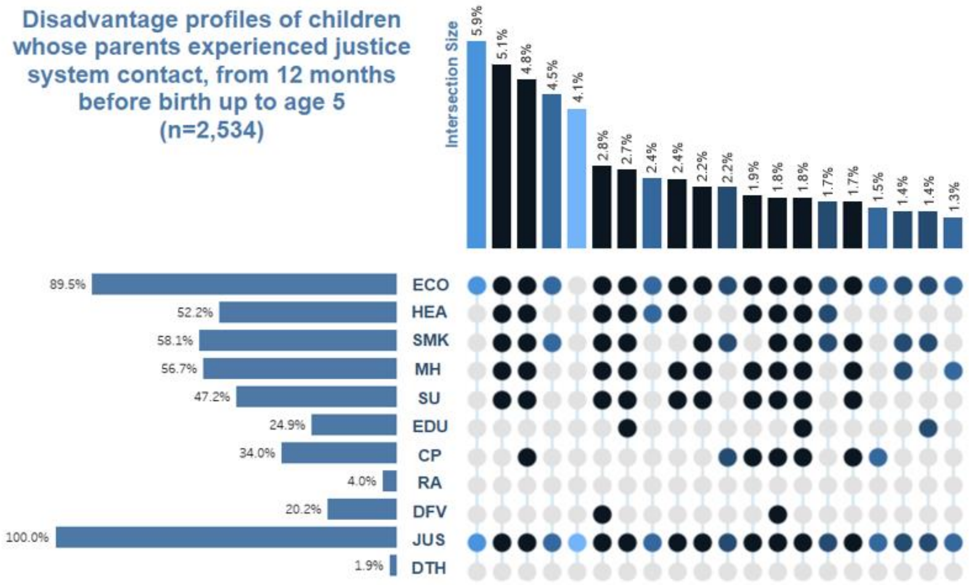

Domain abbreviations: ECO = economic disadvantage, HEA = health disadvantage, SMK = smoking during pregnancy, MH = mental health disadvantage, SU = substance misuse disadvantage, EDU = education disadvantage, CP = child protection disadvantage, RA = access to services disadvantage, DFV = domestic family violence, JUS = justice system contact; DTH = death of parent

Supplementary Figure 5 shows that, among children whose parent experienced justice system contact, the majority (89.5%) experienced economic disadvantage, with the two-domain co-occurrence of economic disadvantage and justice system contact being the most common disadvantage-combination experienced (5.9%). This was followed by the six-domain combination of economic, health, smoking during pregnancy, mental health, substance misuse and justice system contact being the most common disadvantage-combination experienced (5.1%).
